# Racial/Ethnic Disparities in COVID-19 Hospital Admissions

**DOI:** 10.1101/2020.07.12.20152017

**Authors:** Violeta Álvarez Retamales, Oswaldo Madrid Suarez, Odalys E. Lara-Garcia, Suhayb Ranjha, Ruby Maini, Susan Hingle, Vidya Sundareshan, Robert L. Robinson

**Affiliations:** Department of Internal Medicine, Southern Illinois University School of Medicine, Springfield IL; Universidad Central de Venezuela, Luis Razetti School of Medicine. Caracas, Venezuela; Division of Infectious Diseases, Southern Illinois University School of Medicine, Springfield IL

## Abstract

**Importance:** COVID-19 has affected millions of people worldwide. Furthermore, with its increasing incidence, more has been learned about the risk factors that can make certain groups more at risk of contracting the disease or have worse outcomes. We aim to identify any discrepancy in the hospitalization rate by race/ethnicity of patients who tested positive for COVID-19, and through this, analyze the risks of these groups in an effort to call out for attention to the circumstances that make them more vulnerable and susceptible to disease.

**Observations:** Analysis indicates that patients identified as non-Hispanic White and Asian/Pacific Islander in hospital admission data are underrepresented in COVID-19 admissions. Patients identified as non-Hispanic Black, Hispanic/Latino, and American Indian have a disproportionate burden of hospital admissions, suggesting an increased risk of more severe disease.

**Conclusions and Relevance:** There is a disproportionate rate of COVID-19 hospitalizations found among non-Hispanic Blacks. Further investigation is imperative to identify and remediate the reason(s) for increased vulnerability to COVID-19 infections requiring hospital admission. These efforts would likely reduce the COVID-19 morbidity and mortality in the non-Hispanic Black population.

## Introduction

Severe acute respiratory syndrome coronavirus-2 (SARS-CoV-2) has led to a global pandemic manifested as coronavirus disease 2019 (COVID-19) affecting more than eight million people, with its most severe presentation being acute respiratory distress syndrome leading to severe complications and death [1]. With the increment in COVID-19 incidence, more is being learned about which individuals and groups experience the most complications. Researchers have emphasized that older age, male sex, hypertension, diabetes, obesity, concomitant cardiovascular diseases (including coronary artery disease and heart failure), and myocardial injury are significant risk factors associated with worse outcomes [2]. With these factors set in mind, we must acknowledge that racial/ethnic minority populations have a disproportionate burden of underlying comorbidities like diabetes, kidney disease, and cardiovascular disease[3]. Furthermore, certain minority groups have a higher infant mortality rate along with a lower life expectancy at birth, and both influenced in a multifactorial manner[4].

Moreover, the majority of U.S. adults are unaware that racial inequities in health exist [5]. The underappreciated segregation contributes to racial disparities, which can fluctuate between higher rates of underlying disease risk factors and a long-standing lack of access to health care, lack of cultural competency, and adverse social determinants of health [6]. It is worth saying that the problem of segregation is not residing among persons of the same race, but the clustering of social disadvantage and systematic disinvestment in marginalized communities [7]. Racial/ethnic minorities are more likely to live in crowded conditions with particulate matter air pollution, a significant risk factor for cardiopulmonary mortality [8]. In the end, greater exposure to and clustering of stressors contributes to the earlier onset of multiple chronic conditions[7].

Hospital admission criteria are based on medical necessity, which is a subjective and vague term [9]. Yet, could these identified risks change the hospitalization rate of patients who tested positive for COVID-19? There has not been any study or retrospective data showing any racial/ethnic classification of COVID-19 related hospitalizations. Our observational study aims to investigate the rate of racial/ethnic groups of COVID-19 related admissions and identify any possible tendency.

Concern has been raised about the disproportionate impact of COVID-19 on minority populations in the USA [3]. This study aims to use publicly reported data to investigate if racially disparate rates of COVID-19 related hospital admissions can be demonstrated in the USA. This information could be used towards preventive health medicine and development of policies to obtain data on social determinants of health in order to further improve the financial implications associated with COVID-19 complications.

## Methods

Population data were obtained from the U.S. Census Bureau Quick Facts for population estimates as of July 1, 2019 (the most recent available). This data includes an estimate of the overall population and proportions of the population by race. This information was used to calculate the population for each Census-reported racial category as of July 1, 2019 [10].

The U.S. COVID-19 hospital admission data were obtained from the CDC COVID-NET website on June 11, 2020 [11]. This database contains only nationwide counts of COVID-19 hospital admissions by race. All patients with a documented race were included in the analysis. No demographic, severity of illness or location data is included in the COVID-NET dataset.

Statistical analyses were performed using SPSS version 25 (SPSS Inc., Chicago, IL, USA). A p-value of 0.05 was chosen for statistical significance. The potential of the disparate impact of COVID-19 hospital admissions by race was tested with the Chi-square test.

Institutional review board review for this study was obtained from the Springfield Committee for Research Involving Human Subjects. This study was determined not to meet the criteria for research involving human subjects according to 45 CFR 46.101 and 45 CFR 46.102.

## Results

The population as per July 1, 2019 in the USA was estimated at 328,239,523. The demographic analysis allows us to evaluate the racial composition of the U.S. population which is detailed as the following: non-Hispanic White 60.4%, Hispanic or Latino 18.3%, Black or African American 13.4%, American Indian and Alaska Native 1.3%, Asian, Native Hawaiian and other Pacific Islander 6% (Table 1). While for June 11, 2020, the patients hospitalized for COVID-19 infection was 21,221. The racial composition of these hospitalized patients is reported as follows: non-Hispanic White 38%, non-Hispanic Black 36%, Hispanic or Latino 19%, American Indian/Alaska Native 1.6%, Asian, Native Hawaiian and other Pacific Islander 5%.

**Table 1.** U.S. Population estimates demographic data for July 1, 2019

| **Census Reported Race/Ethnicity** | **Population %** |
| --- | --- |
| **Non-Hispanic White** | 60.4% |
| **Hispanic or Latino** | 18.3% |
| **Non-Hispanic Black** | 13.4% |
| **Asian/Pacific Islander** | 6% |
| **American Indian/Alaska Native** | 1.3% |

Analysis indicates that patients reported as non-Hispanic White (60% of the population, 38% of admissions, p >0.001) and Asian/Pacific Islander (6% of the population, 5% of admissions, p>0.001) are underrepresented in COVID-19 admissions (Table 2). Patients reported as non-Hispanic Black (13% of the population, 36% of admissions, p>0.001), Hispanic (18% of the population, 19% of admissions, p>0.001), and American Indian (1.3% of the population, 1.6% of admissions, p >0.001) have a disparate burden of hospital admissions.

**Table 2.** US COVID-19 Hospital Admissions by Reported Race/Ethnicity

| **Reported Race/Ethnicity** | **Hospital**<br>**Admissions** | **Population**<br>**%** | **COVID-19**<br>**%** | **P-value** |
| --- | --- | --- | --- | --- |
| **Non-Hispanic White** | 7530 | 60.4% | 38% | < 0.001 |
| **Hispanic/Latino** | 3797 | 18.3% | 19% | < 0.001 |
| **Non-Hispanic Black** | 7092 | 13.4% | 36% | < 0.001 |
| **Asian/Pacific Islander** | 985 | 6% | 5% | < 0.001 |
| **American Indian/Alaska Native** | 320 | 1.3% | 1.6% | < 0.001 |

The proportion of non-Hispanic Black patients admitted (36%) is significantly higher than the percentage of the general population (13%), suggesting a significant race-based health disparity or vulnerability to disease (Figure 1).

**Figure 1.**
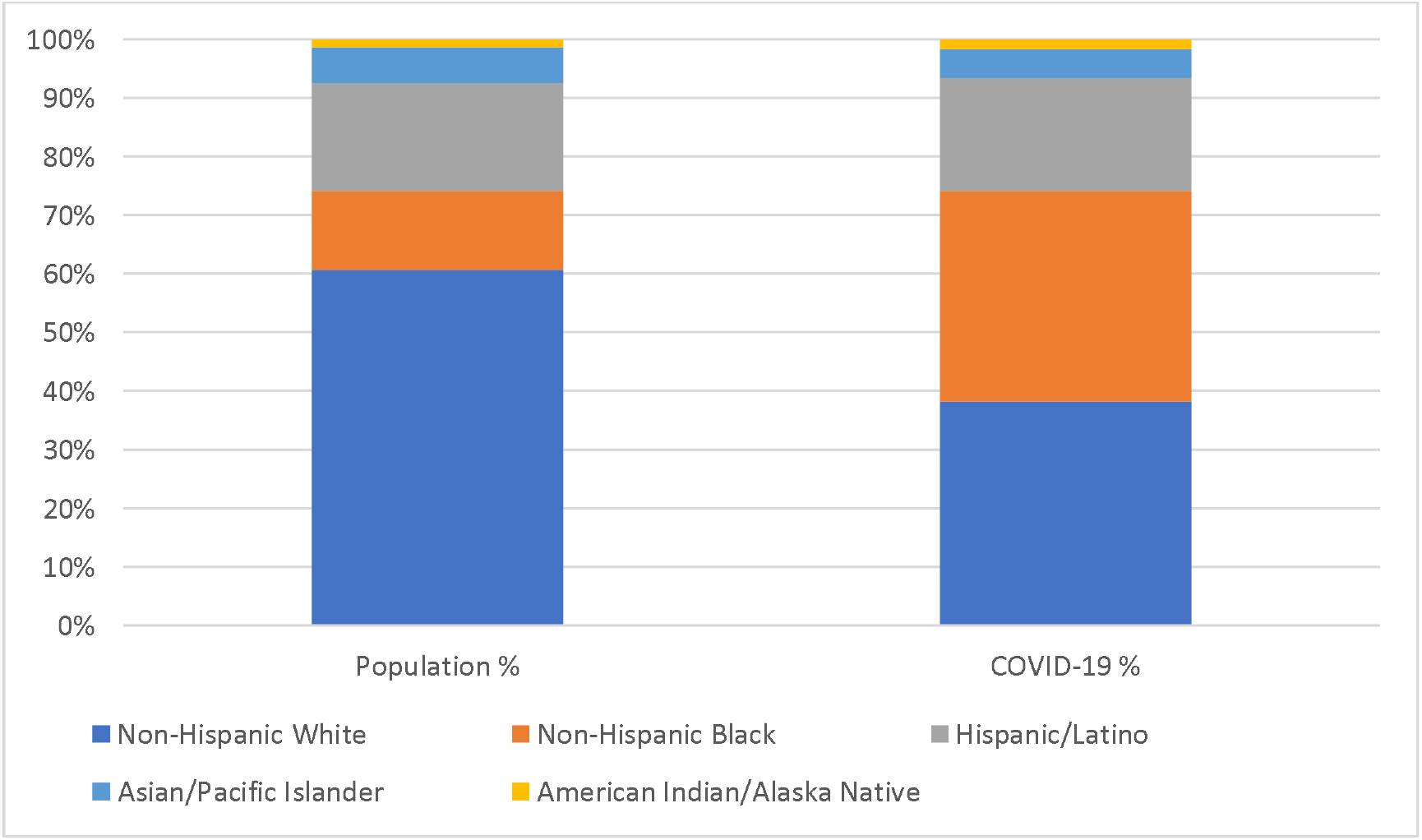
U.S. Population Data and COVID-19 Hospitalization Data stratified by Race/ethnicity.

## Discussion

Based on demographic analysis of the United States population by July 2019, we find that the racial composition is reported to be non-Hispanic White, followed by Hispanic/Latino, non-Hispanic Black, Asians, and Pacific Islanders, and Native Americans (Table 1). After reviewing these results, we could expect a similar racial distribution to be seen in COVID-19 hospitalized patients; however, this is not the case. The racial distribution of hospitalizations shows that despite non-Hispanic Whites representing the majority of COVID-19 admissions, they are closely followed by non-Hispanic Blacks. These findings demonstrate an apparent disproportion of hospitalizations in non-Hispanic Blacks when compared to their overall demographic representation. This data correlates with recently released data from Medicare claims, which reveal that non-Hispanic Black Americans were nearly four times more likely than non-Hispanic White Americans to be admitted to the hospital [12]. Other racial groups such as Asians, Latinos, and Native Americans, have hospitalization rates proportionate to their demographic distributions.

Non-Hispanic Black individuals carry significantly higher comorbidities, are at higher risk for cancer, peripheral arterial disease, hypertension, cardiovascular disease, and diabetes mellitus[13]. Diseases which are biological risk factors for severe outcomes with COVID-19 infection, raising their need for hospital admission [10]. Moreover, in 2017, the difference between the group with the highest (Hispanic) and lowest (non-Hispanic Black) life expectancy at birth was 6.9 years while the infant mortality rate was 170% higher among infants of non-Hispanic black women than among infants of Asian or Pacific Islander women; both revealing the higher mortality rate and worse health status of the non-Hispanic Black population[4].

Nevertheless, it can also be explained by the social circumstances of their everyday life, some of which are also identified by the CDC. Socioeconomic status plays a role in disease outcomes; thus, we should consider it when discussing the hospitalization rate [6]. Social distancing is the most effective strategy to reduce COVID-19 infection, but herein lies a vexing challenge [2]. The ability to isolate in a safe home, work remotely with full digital access, and sustain monthly income are all privileges not all individuals possess [2]. Minority groups are also more likely to be essential workers, making social distancing even more challenging [7].

Furthermore, the convenience of having sick days as part of employment is not feasible for all. Another factor that is associated with socioeconomic status is the ability to obtain health insurance. According to data shared by the CDC, in 2018, 15.2% of non-Hispanic Black adults aged 18–64 lacked health insurance, which is higher than the national average of 9.4% [14].

In the United States, Black, Hispanic/Latino/Latina, and dark-skinned people face disparities in access to health care, the quality of care received, and health outcomes. It is concerning how health care providers’ attitudes and behaviors have been identified as one of many factors that contribute to health disparities. Studies from 2015 and 2017 reported that most health care providers appear to have an implicit bias in terms of positive attitudes toward Whites and negative attitudes toward people of color[7,15]. This inequality in care has also been present during this pandemic as a recent report based on billing data for COVID-19 testing from several states revealed that non-Hispanic Black and Hispanic patients with symptoms such as cough and fever were less likely than white individuals with the same symptoms to be given a test showing yet another barrier that the Black community faces [7].

Taking all these factors into consideration, it is possible that non-Hispanic Black individuals avoid medical care, whether due to physician’s mistrust as victims of their implicit bias or fear of financial repercussion, until their condition worsens and their hospitalization is considered a medical necessity. At the same time, the sole presence of comorbidities for which non-Hispanic Black individuals have a high risk can also increase the medical necessity for hospital admission for COVID-19 infection. The factors that correlate with the lower rate of White individuals admitted for COVID-19 infection are a lower uninsured rate, fewer health barriers, and a higher socioeconomic status than racial/ethnic minorities.

There are many limitations to this study. We assume that the patients admitted met hospitalization criteria based on medical necessity. Yet, we are unable to corroborate this as we are not able to incorporate the population symptoms or severity of illness. At the same time, we can establish the demographics of the admitted population. Still, we cannot identify specific determinants to these results as we are not able to prove it is solely due to Health disparities. We are yet to determine if there is a population protected from COVID-19 due to their race/ethnicity, which could also impact their hospitalization.

## Conclusion

COVID-19 has put the spotlight on apparent social differences among races. A disproportionate rate of COVID-19 hospitalizations found among non-Hispanic Blacks is an example of this. Utilization of community health workers to provide timely and culturally sensitive information along with widespread testing and early provision of personal protective equipment to essential workers such as store clerks; are actions that we could argue may have prevented such a high influx of hospitalizations.

This manuscript aims to report this data and present differences to call out for attention to these groups that are unequally affected by this pandemic and their day-to-day social circumstances that make them more vulnerable and susceptible to disease. Our intention is that future efforts are targeted towards addressing these circumstances.

## Data Availability

Analyzing existing population samples or data set which are recorded by the investigator without identifiers from COVID-NET data set and the U.S. Census Bureau Quick Facts.

https://gis.cdc.gov/grasp/COVIDNet/COVID19_5.html

https://www.census.gov/quickfacts/fact/table/US/PST045219

